# Improving the Prediction of Unplanned 30-day Cancer Readmissions Using Social Determinants of Health: A Geocoding-based Approach

**DOI:** 10.1101/2025.08.31.25334806

**Authors:** Shwetha Bindhu, Tzu-Chun Wu, Hanniel Shih, Himaja Chintalapalli, Hexuan Liu, Anjanette Wells, Caroline F. Morrison, Wei-Wen Hsu, Danny T.Y. Wu

**Affiliations:** School of Medicine, Case Western Reserve University, Cleveland, OH, United States; Department of Biostatistics, Health Informatics, and Data Sciences, College of Medicine, University of Cincinnati, Cincinnati, OH, United States; School of Information and Library Science, University of North Carolina, Chapel Hill, NC, United States

## Abstract

Unplanned cancer readmissions present a significant burden on patients and hospitals. Current predictive models often overlook socioeconomic factors such as social determinants of health (SDoH), which have the potential to improve prediction performance, as measured by the Area Under the Receiver Operating Characteristic (AUROC) and the Precision-Recall Curve (AUPRC). To investigate this, the present study developed predictive models using cancer readmission data from a large health system in Hamilton County, OH. The models incorporated geocoding-based SDoH along with clustering techniques and compared machine learning (ML) and deep learning (DL) algorithms. Overall, models, regardless of algorithm type, not using SDoH variables had higher AUROC and AUPRCs. The best-performing ML and DL models are comparable (AUROC = 0.7605 for ML; AUROC = 0.7585 for DL). However, when top-performing models were evaluated across certain organ and system cancers, using SDoH and clustering techniques significantly improved model performance. This was most notable for cancers of the skin, subcutaneous tissue, and breast with improvements of 8.20% in AUROC and 11.04% in AUPRC. For all cancer patient cases, utilizing individualized SDoH information extracted from clinical notes was recommended for future studies.

## Introduction

Cancer diagnoses can be devastating medical news for patients and their families. Unplanned cancer readmissions add to the substantial burdens of treatment and disease management by increasing incurred costs and mortality(1,2). These readmissions also happen more frequently among uninsured and lower-income patients, which worsens existing disparities in cancer treatment(2–5). Predicting patients risk of unplanned readmissions can mitigate these disparities. Current methods use electronic health records (EHR) and ML algorithms to predict unplanned readmissions. Models are often trained on clinical and administrative data, including variables such as demographic information and patient medical history(6).

However, models often do not account for the impact of social determinants of health (SDoH) and health inequities, which have been found to increase all-cause readmission risk as social needs increase (7). One study showed that neighborhood factors and crime index contributed to more unplanned cancer readmissions(8). While the authors found that their model omitting SDoH factors performed better (Area Under the Receiver Operating Characteristic (AUROC) = 0.80) than the model incorporating SDoH (AUROC = 0.78), having the context of a patient’s surrounding environment provides a more holistic picture of patient risk. Another study focused on sepsis readmissions found that using SDoH criteria could increase the predictive model performance(9). Moreover, readmissions rates alone are an imperfect indicator of health care needs, and more frequent utilization of emergency healthcare services may underestimate access barriers in routine disease management(4). Therefore, including SDoH to stratify patient readmission risk may provide crucial context that can improve the relevance of predictive models, especially for marginalized and underserved communities.

One plausible way to collect SDoH variables is geocoding, which assigns coordinates to an address and uses census data to infer SDoH. During the past two decades, geocoding has been used to understand and map spatial health inequities(10,11). The Decentralized Geomarker Assessment for Multi-Site Studies (DeGAUSS) project geocoded addresses to census tract, added community-level geomarker information such as median household income, and deidentified the information by removing the address and location information(12). This method increased the number of geocoded results and subject inclusion. Using geocoding in predictive modeling could help identify and visualize socioeconomic disparities in cancer readmission because it includes patient’s neighborhood and built environments as prominent readmission risk factors(8).

Recent advances in artificial intelligence offer another opportunity to improve model performance. Meanwhile, EHR data can be complex and difficult to analyze with ML models alone. DL algorithms incorporate layered structures that could capture nuanced information that ML algorithms may miss, possibly making them better at predicting readmissions(13). In a comparison of ML and DL models for predicting readmissions for asthma and chronic obstructive pulmonary disease (COPD), the DL model showed better sensitivity and specificity in predicting high-risk patient readmission(14). The higher model performance can result in cost savings by minimizing unnecessary readmissions and improving resource utilization(15).

This study aims to explore strategies for enhancing the prediction of unplanned cancer readmissions and address the following research questions: 1) Can incorporating geocoding-based SDoH factors improve ML model performance? 2) Does clustering patients based on their SDoH profiles enhance model performance? 3) Will DL models outperform traditional ML algorithms in predicting readmissions? 4) Are there improvements in prediction accuracy across different major disease types?

## Methods

### Study Setting

The study was a collaboration between the University of Cincinnati Cancer Center (UCCC) and the UC Center for Health Informatics (CHI). UCCC aims to address cancer mortality in the Greater Cincinnati area by providing patient-specific care involving multidisciplinary teams and specialized treatment plans. The UCCC offers treatment for a broad array of cancer subtypes and emphasizes preventive care through cancer screening programs.

### Previous Work and Present Study

Developing the ML models followed methods outlined in our previously published paper on predicting unplanned cancer readmission(6). In this paper, the 3-year dataset (N=156,816) before pandemic from 2017 to 2019. The dataset was split into training and testing sets based on the admission dates (first 2-year dataset for training), and the values of the outcome variable were updated based on the latest definition of readmissions at UC Health. Five models were trained using ML classifiers, including Gradient Boosting Machine (GBM), decision tree (DT), random forest (RF), Ridge logistic regression (LR), and Support Vector Machines with Linear Kernal (linear SVM). The AUROC and the Areas Under the Precision and Recall Curve (AUPRC) were used to determine model performance. The GBM model achieved the highest AUROC on the testing set. The dataset was extracted by the Center for Health Informatics (CHI) for research purposes from our institution’s electronic health records on August 9, 2023. Only the corresponding author (DW) and the lead data analyst (TW) had access to the identifiable patient information in the initial data dataset on the secure server of CHI, which was de-identified for all downstream analyses. The study was reviewed and approved by the Institutional Review Board (IRB #2021-0003).

### SDoH Variables

The SDoH variables came from two sources: 1) the Decentralized Geomarker Assessment for Multi-Site Studies (DeGAUSS) pipeline(12) and 2) the National Cancer Index (NCI). The DeGAUSS variables address key SDoH, such as deprivation indices, household income, and food security. The NCI dataset was based on the 2010 U.S. census tracts, which captured sociodemographic characteristics, such as race and education level.

### Clustering Method

Consensus clustering was used to create SDoH profiles, which is a robust method to enhance the reliability of clustering results by integrating multiple clustering outcomes into a single consensus solution(16). This improves the stability and interpretability of clusters, as it mitigates the variability seen in a single clustering algorithm or parameter setting. Consensus clustering results were first obtained from training data and used to predict clusters in this data by assigning each test sample to the nearest cluster center of SDoH identified during the training phase.

The analysis was performed using the R package ‘ConsensusClusterPlus’ version 1.68.0 in R version 4.4.0, utilizing K-means clustering with 1,000 replications on 80% subsampling. The optimal number of clusters was determined by the within-cluster consensus scores and the matrix heat map. In the testing data set, each data point was assigned to the cluster with the nearest cluster centroid.

### Deep Learning Model

Additionally, a Deep Neural Network (DNN) consisting of three Feed Forward Layers with residual was implemented, using the Sigmoid Linear Unit (SiLU) as the activation function for hidden layers, for better convergence and performance, and Sigmoid for the final classification layer (17–19). Residual connections have been shown to improve performance for tabular data in for DNNs (20,21). Dropout and Batch Normalization were implemented between hidden layers for regularization and to reduce over-fitting(22,23). The model was trained using the AdamW optimizer for 50 epochs, where the learning rate is adjusted with an exponential warm-up of 20 epochs and an exponential decrease afterward, and Binary Cross Entropy as the loss function(24–26).

### Performance Comparison

The dataset contains 24 major diagnostic categories (MDCs) across broad disease types and affected organ systems To compare model performance within each MDC, the DeLong test was used to assess the AUROCs, while the permutation test was employed to evaluate the AUPRCs between models. A significance level of 0.05 was set for determining statistical significance based on the p-value.

To answer our research questions, 16 models with various settings were generated. The settings resulted from two types of algorithms (ML or DL), 4 ways to include SDoH variables (None, DeGuass Only, NCI Only, or both), and whether patients were grouped by SDoH profiles before training the models. Again, the model performance was determined using AUROC and AUPRC. AUPRC was considered because the dataset was imbalanced (the positive rate was 25.29%). Once the top-performing models are selected, the performances of each MDC are compared. It is hypothesized that each of the strategies or within certain MDCs, namely, DL, SDoH profiling, and SDoH variables, will improve the prediction performance of unplanned cancer readmissions.

## Results

Based on the consensus clustering results, three clusters were selected for model comparison. As shown in **Table 1**, the ML models (#1-8) had greater AUROCs, ranging between 0.7574 to 0.7605 while DL models (#9-16) had AUROCs ranging from 0.7435 to 0.7592. Moreover, the baseline model (#1) using ML without SDoH profiling nor SDoH variables performed the best overall with an AUROC of 0.7605. The models using SDoH profiling (#5-8, #13-16), regardless of algorithm type, had decreased AUROC and AUPRC. Regarding the use of geocoding-based SDoH variables, the models without such SDoH variables had higher AUROC and AUPRC than those with such variables (#1 versus #2-4; #5 versus #6-8, #9 versus #10-12, #13 versus #14-16).

**Table 1.** Performance and breakdown and of the ML and DL models, including various variables each incorporates.

| Model | Algorithm | SDoH Profile | Geocoding-based SDoH Variable | AUROC | AUPRC |
| --- | --- | --- | --- | --- | --- |
| 1 | ML | No | No | **0.7605** | 0.5437 |
| 2 | ML | No | Yes – DeGAUSS Only | **0.7601** | 0.5439 |
| 3 | ML | No | Yes – NCI Only | **0.7600** | 0.5419 |
| 4 | ML | No | Yes – Both | 0.7591 | 0.5405 |
| 5 | ML | Yes Consensus | No | 0.7580 | 0.5340 |
| 6 | ML | Yes Consensus | Yes – DeGAUSS Only | 0.7574 | 0.5318 |
| 7 | ML | Yes Consensus | Yes – NCI Only | 0.7578 | 0.5337 |
| 8 | ML | Yes Consensus | Yes – Both | 0.7574 | 0.5307 |
| 9 | DL | No | No | 0.7585 | **0.5508** |
| 10 | DL | No | Yes – DeGAUSS Only | 0.7592 | **0.5499** |
| 11 | DL | No | Yes – NCI Only | 0.7564 | 0.5422 |
| 12 | DL | No | Yes – Both | 0.7566 | **0.5446** |
| 13 | DL | Yes Consensus | No | 0.7505 | 0.5335 |
| 14 | DL | Yes Consensus | Yes – DeGAUSS Only | 0.7494 | 0.5299 |
| 15 | DL | Yes Consensus | Yes – NCI Only | 0.7435 | 0.5150 |

Moreover, **Table 2** presents a ranking of various health-related features across different models. The features include factors such as emergency department visits, discharge dispositions, oncology treatment, and pregnancy. Each feature is assigned a ranking within each model, indicating its importance in the model’s predictive capability. The top three features of the best model (#1) were “hospital class: Hosp OP Surg/Ambulatory”, “number of emergency department visits within past 6 months”, “discharge disposition: home or self-care without home care services”. When reviewing all top 10 features from all models, only two were directly correlated to a cancer diagnosis: “oncology treatment” and “myeloproliferative differential diagnoses (poorly differentiated neoplasms).” Additionally, the SDoH variables and “age during admission” were not considered as one of the top 10 features in any model. However, as previously discussed, repeat emergency admissions may suggest several health care needs are unmet (4).

**Table 2.**
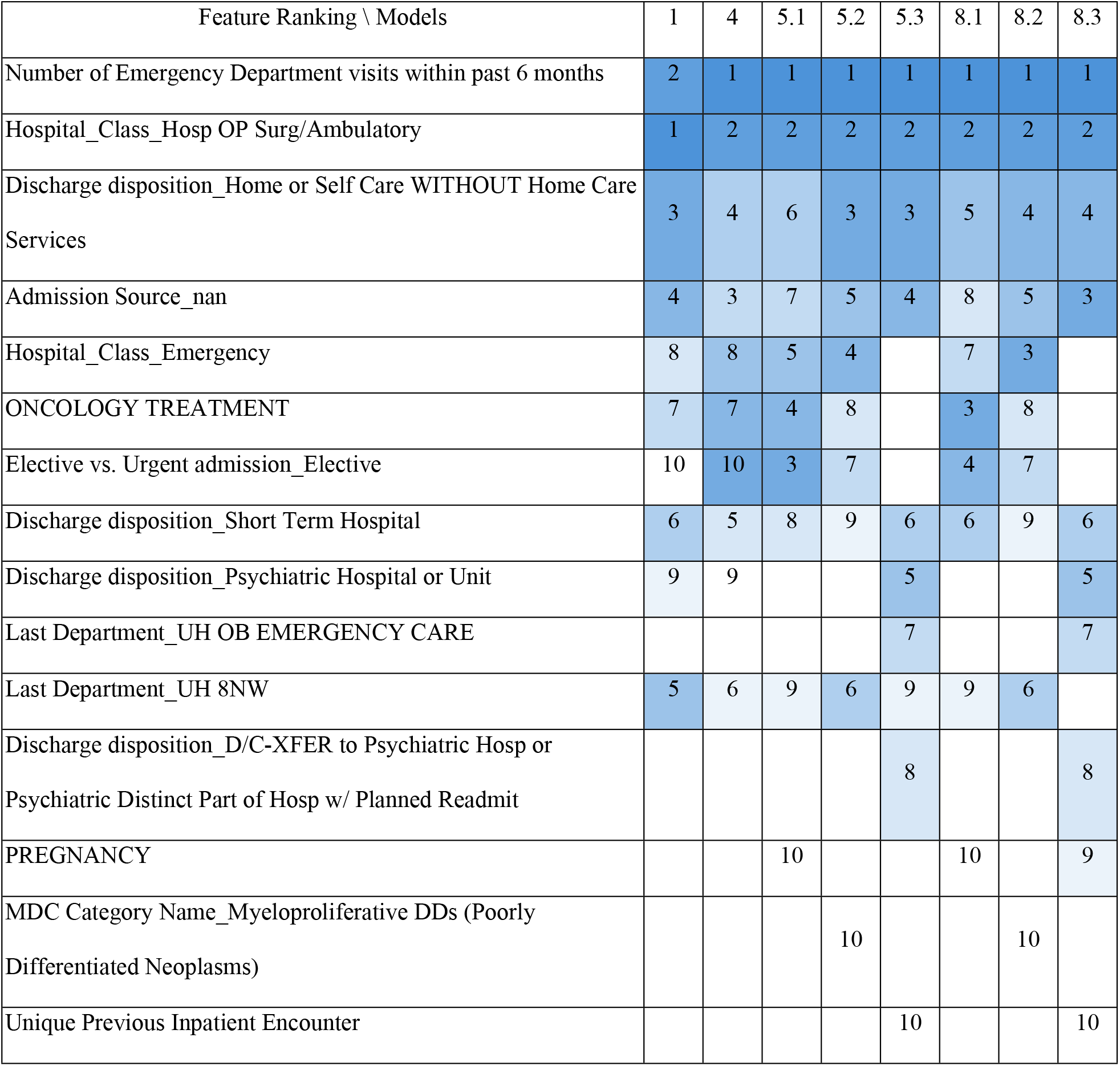
Feature importance ranking of machine learning models. 5.1 means the first cluster of model 5.

| Feature Ranking \ Models | 1 | 4 | 5.1 | 5.2 | 5.3 | 8.1 | 8.2 | 8.3 |
| --- | --- | --- | --- | --- | --- | --- | --- | --- |
| Number of Emergency Department visits within past 6 months | 2 | 1 | 1 | 1 | 1 | 1 | 1 | 1 |
| Hospital_Class_Hosp OP Surg/Ambulatory | 1 | 2 | 2 | 2 | 2 | 2 | 2 | 2 |
| Discharge disposition_Home or Self Care WITHOUT Home Care Services | 3 | 4 | 6 | 3 | 3 | 5 | 4 | 4 |
| Admission Source_nan | 4 | 3 | 7 | 5 | 4 | 8 | 5 | 3 |
| Hospital_Class_Emergency | 8 | 8 | 5 | 4 |  | 7 | 3 |  |
| ONCOLOGY TREATMENT | 7 | 7 | 4 | 8 |  | 3 | 8 |  |
| Elective vs. Urgent admission_Elective | 10 | 10 | 3 | 7 |  | 4 | 7 |  |
| Discharge disposition_Short Term Hospital | 6 | 5 | 8 | 9 | 6 | 6 | 9 | 6 |
| Discharge disposition_Psychiatric Hospital or Unit | 9 | 9 |  |  | 5 |  |  | 5 |
| Last Department_UH OB EMERGENCY CARE |  |  |  |  | 7 |  |  | 7 |
| Last Department_UH 8NW | 5 | 6 | 9 | 6 | 9 | 9 | 6 |  |
| Discharge disposition_D/C-XFER to Psychiatric Hosp or Psychiatric Distinct Part of Hosp w/ Planned Readmit |  |  |  |  | 8 |  |  | 8 |
| PREGNANCY |  |  | 10 |  |  | 10 |  | 9 |
| MDC Category Name_Myeloproliferative DDs (Poorly Differentiated Neoplasms) |  |  |  | 10 |  |  | 10 |  |
| Unique Previous Inpatient Encounter |  |  |  |  | 10 |  |  | 10 |

Further analysis of the top-performing model, both with and without SDoH (#4 and #1), as well as their corresponding models with SDoH profiling (#8 and #5), is shown in **Table 3**. The models with at least one significant improvement in AUROC or AUPRC, with positive improvements when compared to the baseline model (#1), was evaluated across the following MDCs: Hepatobiliary System and Pancreas, The Skin, Subcutaneous Tissue and Breast, The Blood and Blood-Forming Organs and Immunological Disorders, and Infectious and Parasitic Diseases (Systemic or Unspecified Sites). When applying SDoH variables as features or in profiling, the performance showed significant improvements, especially in AUPRC, with an average increase of 7.53%, ranging from 4.62% to 11.04%. AUROC also showed a compatible improvement, with an average increase of 3.75%, ranging from 0.74% to 8.20%.

**Table 3.**
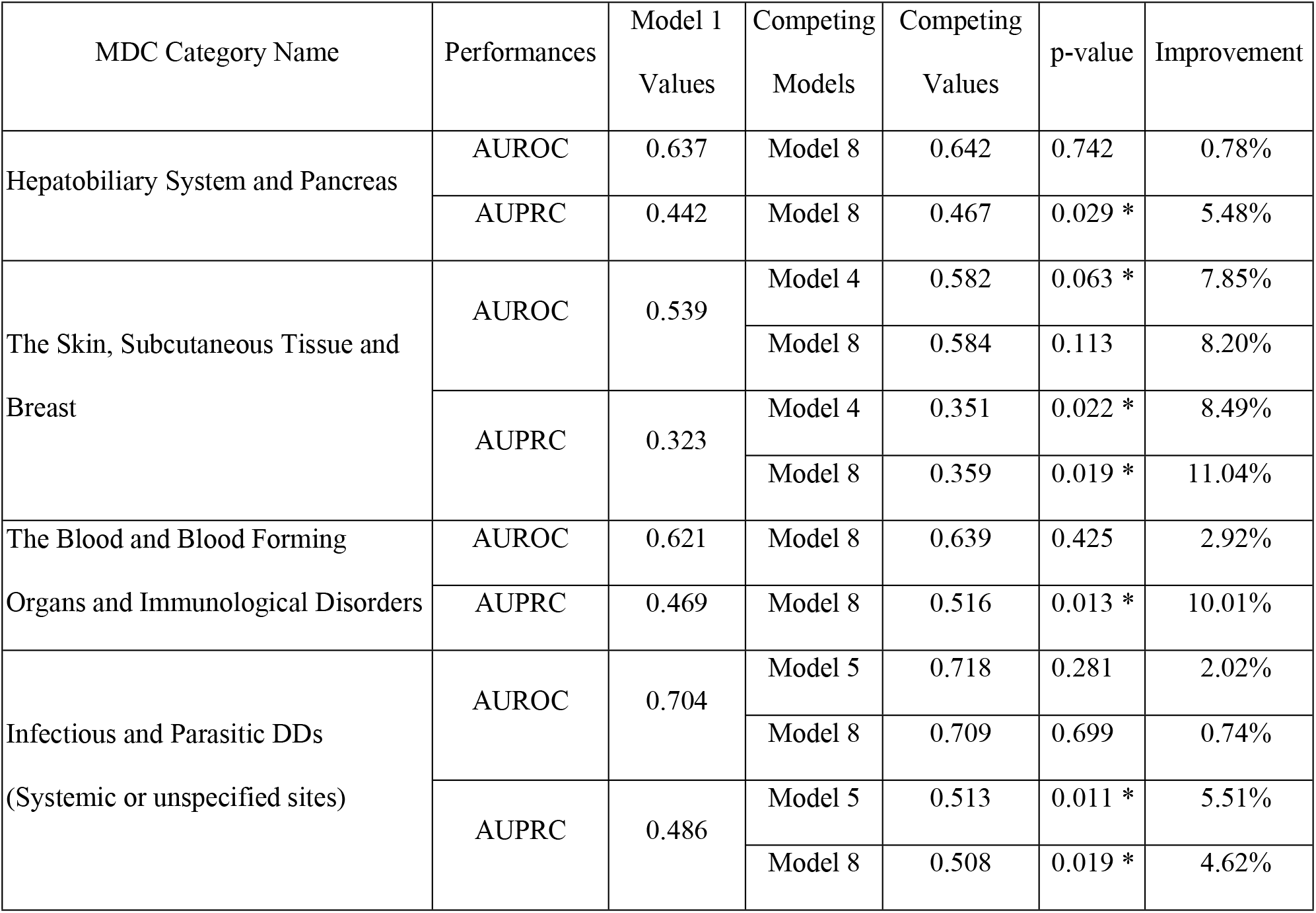
Model improvement is compared with the baseline model. P-values marked with an asterisk indicate that they are lower than the 0.05 significance level.

| MDC Category Name | Performances | Model 1<br>Values | Competing<br>Models | Competing<br>Values | p-value | Improvement |
| --- | --- | --- | --- | --- | --- | --- |
| Hepatobiliary System and Pancreas | AUROC | 0.637 | Model 8 | 0.642 | 0.742 | 0.78% |
|  | AUPRC | 0.442 | Model 8 | 0.467 | 0.029 * | 5.48% |
| The Skin, Subcutaneous Tissue and Breast | AUROC | 0.539 | Model 4 | 0.582 | 0.063 * | 7.85% |
|  |  |  | Model 8 | 0.584 | 0.113 | 8.20% |
|  | AUPRC | 0.323 | Model 4 | 0.351 | 0.022 * | 8.49% |
|  |  |  | Model 8 | 0.359 | 0.019 * | 11.04% |
| The Blood and Blood Forming Organs and Immunological Disorders | AUROC | 0.621 | Model 8 | 0.639 | 0.425 | 2.92% |
|  | AUPRC | 0.469 | Model 8 | 0.516 | 0.013 * | 10.01% |
| Infectious and Parasitic DDs<br>(Systemic or unspecified sites) | AUROC | 0.704 | Model 5 | 0.718 | 0.281 | 2.02% |
|  |  |  | Model 8 | 0.709 | 0.699 | 0.74% |
|  | AUPRC | 0.486 | Model 5 | 0.513 | 0.011 * | 5.51% |
|  |  |  | Model 8 | 0.508 | 0.019 * | 4.62% |

## Discussion

### Key Findings

The present study aimed to assess how incorporating SDoH variables and geocoding may improve model accuracy in predicting unplanned cancer readmissions. Four strategies were examined in the process: adding SDoH variables, constructing SDoH profiling, comparing DL models, and investigating improvement within MDCs. SDoH profiles and geocoding results taken from the previously published DeGAUSS project and NCI variables were used to analyze a dataset composed of patients at UC Health in our previous work. We hypothesized that building SDoH profiles provided a more holistic representation of how socioeconomic factors contribute to patient susceptibility for readmission. Likewise, in comparing the efficacy of ML and DL models in readmission prediction, we anticipated that DL models would be better able to capture these nuances. Lastly, the model’s performance within certain MDCs shows significant improvement. The ML models had greater AUROCs, ranging between 0.7574 and 0.7605 while DL models had AUROCs ranging from 0.7435 and 0.7592. Moreover, the ML model without SDoH profiling, DeGAUSS, or NCI variables performed the best, with an AUROC of 0.7605. Significant improvements were observed in four MDCs with a positive increase in AUROC and a notable enhancement in AUPRC, averaging 3.57% and 7.53%, respectively. These results have valuable implications, as outlined below.

### Implications

Our results showed that the model with both SDoH and profiling can significantly improve the performance of model in certain MDCs instead of all cases. Similarly, the previous study found that their SDoH-based model had a lower AUROC for cancer readmission prediction(8). Another research study also found that including SDoH at individual and census-tract levels in their prediction models did not meaningfully improve prediction accuracy(27). We suspect that geocoding-based SDoH is too general to capture individual risk profiles, especially in populations with large variations in SDoH within census tracts. The census tracts used in the NCI dataset are defined by physical boundaries, which means that population sizes within a single tract can vary from a few to several thousand. For example, in Hamilton County, the site of the present study, the tracts can range from 1,200 to 8,000 individuals and cover unequal geographic areas. While SDoH is important to consider in readmission, geocoding-based SDoH factors may not be granular enough when predicting readmissions. The rankings in **Table 2** suggest that clinical condition acuity has a greater impact on the immediate risk of readmission. We suspect that SDoH factors may have a more longitudinal effect; for example, distance from a healthcare facility may delay care, but a poorly differentiated neoplasm has a worse clinical prognosis and is more likely to necessitate frequent hospital visits. Nevertheless, while SDoH factors were not explicitly listed, the top-ranked features have implications on patient social conditions. A patient who is discharged home without health care services or support can have limitations in the SDoH domains of social and community and healthcare quality. Therefore, individual-level SDoH, such as self-reported factors or those extracted from clinical notes, may be more helpful in readmission prediction. The results also suggest that ML models outperformed DL models, as shown by the slightly higher AUROCs outlined in **Table 1**. However, the DL models had higher AUPRCs. The difference in performance may be due to the imbalanced dataset. Likewise, the underperformance of our models compared to the AUROCs reported by other models in the literature may be due to variations in the dataset population as mentioned above and differences in what variables were used to form them.

### Limitations

There were several limitations to this study. First, this was a single-site study taking place in an urban health center that serves a large population in southeast Ohio and northwest Kentucky. The results may not be generalizable to populations that are rural or serve more homogenous communities and may need to reassessed with datasets that reflect these demographics. Additionally, the data does not include a comparison of before and after the COVID-19 pandemic, which caused noticeable disruption to readmission procedures and emergency medical services. To assess the breadth of this change and what factors may affect readmission risk, an updated analysis is needed with new patient and 2020 Census data. Lastly, this readmission study focused on cancer as the primary diagnosis. Various cancers have different risks for emergencies and unplanned readmissions and are often a more chronically managed disease process. Using SDoH in readmission models for more acute conditions, such as traumatic brain injury or sepsis, may have different implications in model accuracy. Lastly, geocoding-based SDoH approach replied on the census data, which may not reflect the most recent characteristics of the patient populations.

### Future Directions

There are several directions for continuing research. This project provided a baseline for SDoH-based readmission prediction in all cancer types. By narrowing scope and analyzing clinic notes for patients with a specific cancer, such as breast cancer, it may be possible to parse individual SDoH that allows for more accurate and specific predictions. Moreover, using the current study design in predicting readmission risk in more acute conditions may offer insight into how SDoH influences unplanned readmissions in acute versus chronic conditions.

## Conclusion

This study explored the role of geocoding-based SDoH variables and geocoding in predicting unplanned cancer readmissions and found that these factors significantly improve model performance in four MDCs. While ML models outperformed DL models, none surpassed the AUROC benchmark from prior research. The findings suggest that geocoding-based SDoH may be too generalized to capture all-case cancer patient’s risk level, especially in heterogenous populations. Limitations such as the study design and the exclusion of pandemic-related disruptions may have influenced the results. To validate the efficacy of the models and improve their predictive ability, they should be applied on datasets from multiple healthcare settings, including urban and rural, and updated to include post-COVID-19 data. Future research should focus on cancer-specific SDoH analysis and explore the impact of SDoH in predicting readmissions for acute conditions.

## Data Availability

The data were protected by HIPAA and cannot be shared without a data use agreement.

## Acknowledgments

None

## Notes

### Competing Interest Statement

The authors have declared no competing interest.

### Funding Statement

The author(s) received no specific funding for this work.

### Author Declarations

This study was reviewed by the University of Cincinnati Internal Review Board (IRB) and determined as Non-Human Subject Research (#2021-0003).

